# School-based screening and treatment may reduce *P. falciparum* transmission

**DOI:** 10.1101/2020.11.11.20229336

**Authors:** Lauren M. Cohee, Clarissa Valim, Jenna E. Coalson, Andrew Nyambalo, Moses Chilombe, Andrew Ngwira, Andy Bauleni, Karl B. Seydel, Mark L. Wilson, Terrie E. Taylor, Don P. Mathanga, Miriam K. Laufer

## Abstract

In areas where malaria remains entrenched, novel transmission-reducing interventions are essential for malaria elimination. We report the impact of screening-and-treatment of asymptomatic schoolchildren (N=705) on gametocyte - the parasite stage responsible for human-to-mosquito transmission - carriage and use concomitant household-based surveys to predict the potential reduction in transmission in the surrounding community. Among 179 students with gametocyte-containing infections, 84% had positive malaria rapid diagnostic tests. While gametocyte burden remained constant in untreated children, treatment with artemether-lumefantrine reduced the gametocyte prevalence (p<0.0001) from 51.8% to 9.7% and geometric mean gametocyte density (p=0.008) from 0.52 to 0.05 gametocytes/microliter. Based on these estimates, the gametocyte burden in the community could be reduced by 25-55% depending on the season and the measure used to characterize gametocyte carriage. Thus, school-based interventions to treat asymptomatic infections may be a high-yield approach to not only improve the health and education of schoolchildren, but also decrease malaria transmission.

## Introduction

In the last two decades, many regions have made substantial progress toward elimination of malaria. Recently, however, progress has stalled and malaria remains entrenched in some highly endemic areas like Malawi.^1^ In these areas, current interventions (e.g., vector control, universal access to effective testing and treatment, and preventive treatment of high risk populations^2^) may not adequately interrupt human-to-mosquito *Plasmodium falciparum* transmission. Sub-clinical infections in individuals at lower risk of disease often remain untreated, creating a silent reservoir that may perpetuate transmission.^3^ To achieve malaria elimination, interventions must not only prevent and treat disease but also decrease parasite transmission.^4^

School-age children bear an under-appreciated burden of *P. falciparum* infection and may be a key transmission reservoir.^5^ School-age children frequently have higher prevalence of infection than younger children and adults^6–10^ and their infections more often contain gametocytes, the parasite stage required for human-to-mosquito transmission.^8,11^ When bitten, infected school-age children and young children are similarly infectious to mosquitoes.^12^ However, school-age children have larger body surface area and less bed net use, increasing availability for bites.^13–16^ Together these factors suggest school-age children are dominant sources of human-to-mosquito *P. falciparum* transmission.^16–18^ Indeed, expanding community-based seasonal malaria chemoprevention from only young children to include school-age children was associated with a 20% decrease in clinical malaria in older community members who did not receive the intervention.^19^

With rising school enrollments across the malaria-endemic world, malaria interventions could leverage existing school infrastructure. Treatment options include “intermittent preventive treatment” where all students regularly receive antimalarial drugs for parasite clearance and transient prophylaxis, and “screening-and-treatment” where asymptomatic students are screened and only those testing positive receive treatment. Preventive treatment directly decreases infection and gametocyte prevalence, clinical disease, and anemia among students^20,21^, but potential indirect effects on the surrounding community have not been established. While dynamic modeling data suggests the utility of this intervention, only one study has reported the impact of school-based preventive treatment on community-level infection, finding a small but significant decrease in community infection prevalence surrounding intervention schools despite low intervention coverage.^22,23^

We conducted a school-based screen-and-treat cohort study in Malawi concurrent with household-based cross-sectional surveys. Primary school students in four schools were screened for infection using a rapid diagnostic test (RDT). If positive, students were treated with artemether-lumefantrine. Subsequent gametocyte carriage in treated and untreated students was assessed after one, two, and six weeks. Using the age distribution of gametocyte containing infections in the communities surrounding the school, we estimated the potential reduction in prevalence of gametocyte-containing infections and gametocyte density in the community following screening-and-treatment of schoolchildren. If school-based preventive treatment substantially reduces gametocyte burden in the community, this intervention could both improve the health of schoolchildren and contribute more broadly to malaria elimination.

## Results

We enrolled 786 students in school-based cohorts: 405 in the rainy season and 381 in the dry season. Complete follow-up was obtained for 616 students (78% of enrolled students) (Supplemental Figure S1). Among the 705 students with baseline data, the mean age was 10.4 years, 51% were female, and 47% reported sleeping under a net the previous night. The weighted prevalence of RDT-positivity among students at baseline was 42%. An additional 8% of students had infection detected by only PCR. The distribution of infection prevalence varied by school and season (Figure 1).

**Figure 1:**
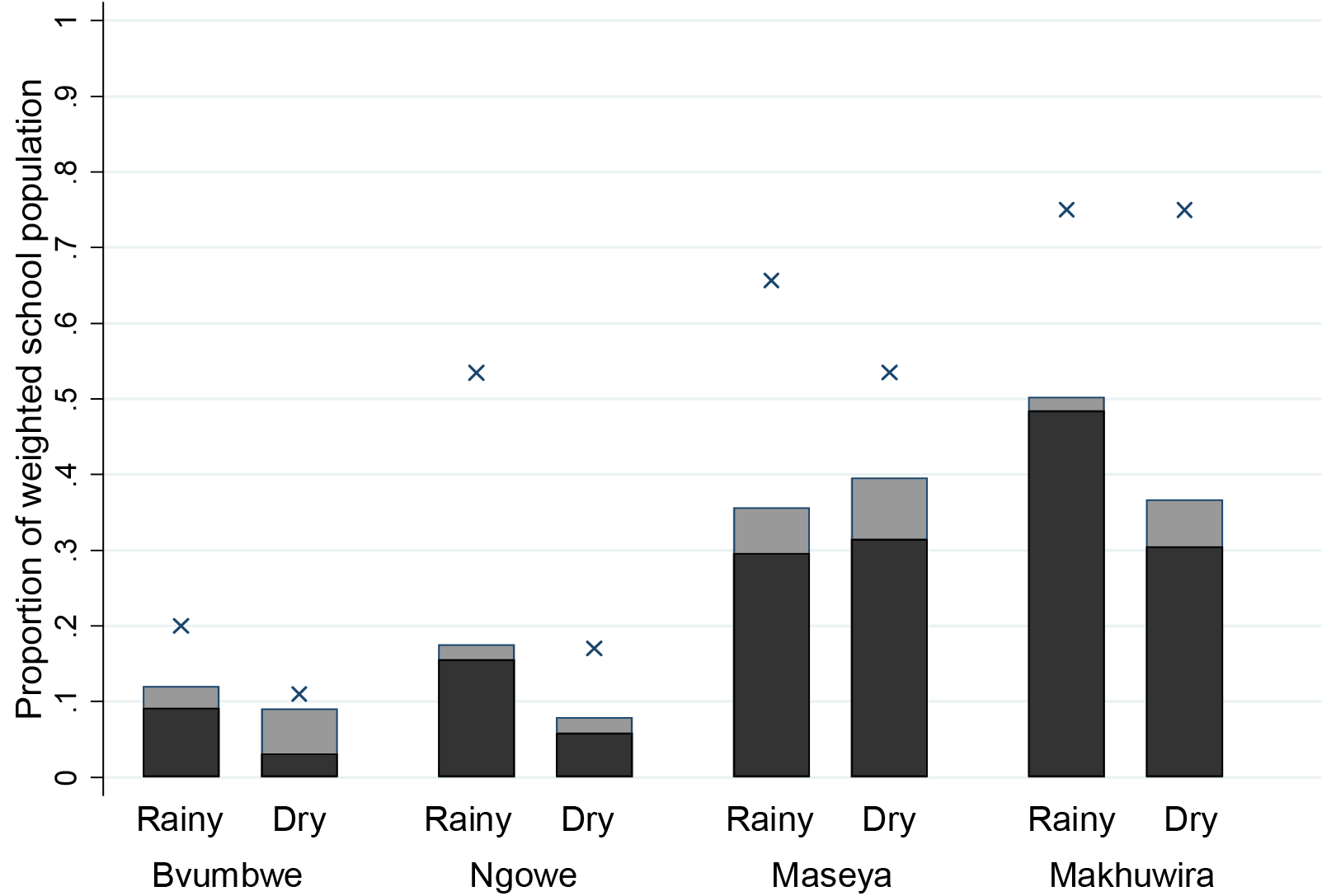
Baseline *P. falciparum* infection prevalence (×) and proportion of students with gametocyte-containing infections detected (black bar) and not detected (grey bar) by rapid diagnostic test in school-based cohorts.

### Determinants of gametocyte burden in school-based cohorts at baseline

Overall, at baseline, 28% of students had gametocyte-containing infections (8-50% across schools and seasons; Figure 1). To disentangle predictors of having an infection containing gametocytes from those of simply having any infection, we evaluated predictors among only students with *P. falciparum* infection. Among the 253 students with *P. falciparum* infection by PCR at baseline, 70% contained gametocytes. Infections were more likely to contain gametocytes in the rainy season and in younger students, although the age association was not statistically significant (Table 1). Associations were comparable in multivariable models with or without non-significant variables.

**Table 1:**
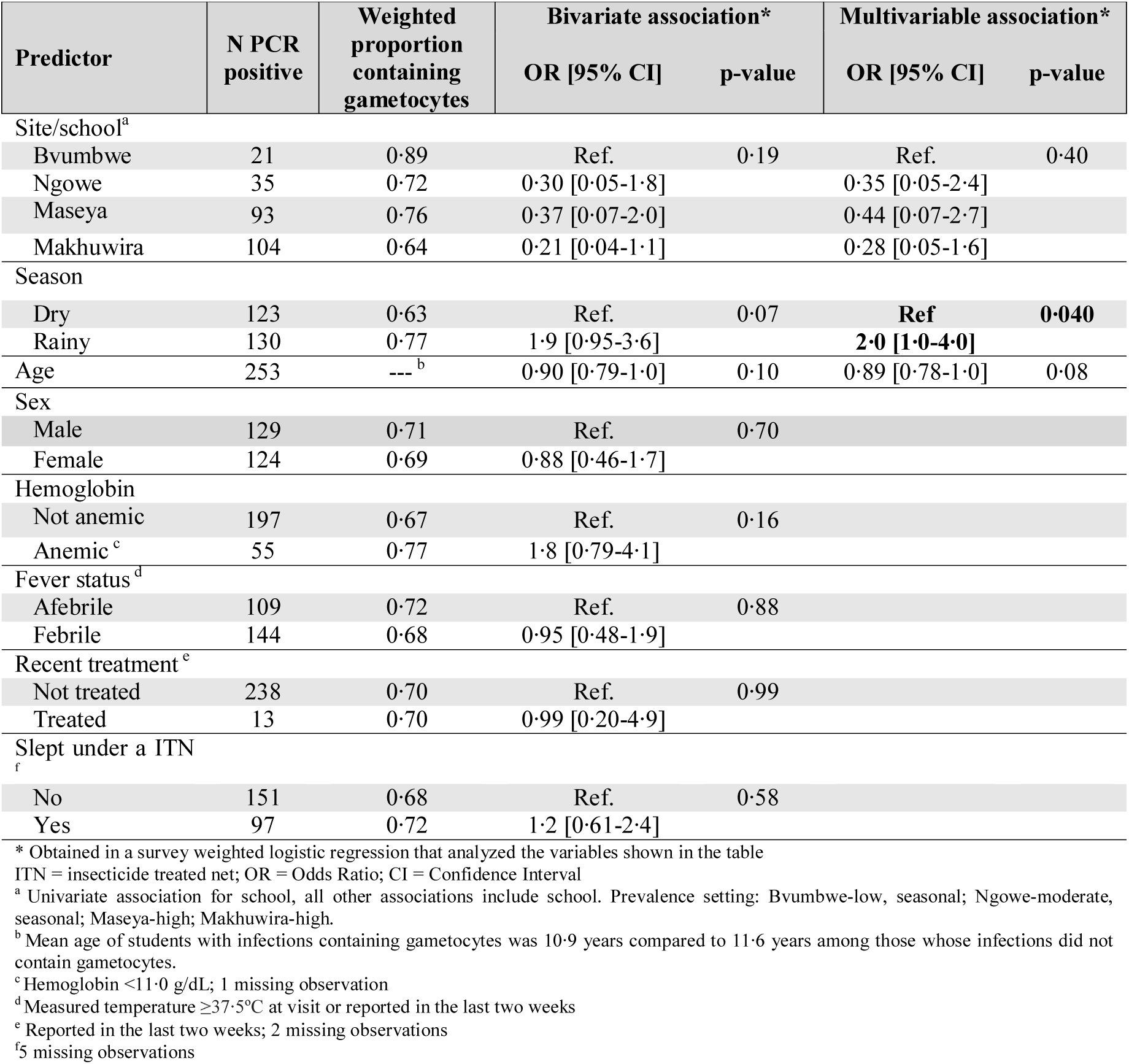
Baseline prevalence and predictors of infections containing gametocytes among students with *P. falciparum* infections in the school-based cohort.

The crude weighted geometric mean gametocyte density among those with gametocytes was 0.34 gametocytes/µl (95% CI: 0.19-0.60 gametocytes/µl; range 0.002-661 gametocytes/µl). Gametocyte density was higher in the rainy season and decreased with age (Table 2). Overall, 5.3% of students had high-density infections (≥10 gametocytes/µl, associated with increased likelihood of transmission^24^) at baseline. Among students with gametocyte-containing infections, 19% had high density infections. In addition to season and age, odds of high density gametocyte infection were significantly higher in children with fever and in the lowest prevalence school (Bvumbwe; Supplemental Table S3).

**Table 2:** Predictors of baseline gametocyte density among infections containing gametocytes in the school cohort.

| Predictor | N<br>gametocyte<br>positive | Bivariate associations* |  | Multivariable associations* |  |
| --- | --- | --- | --- | --- | --- |
|  |  | % Difference**<br>[95% CI] | p-value | % Difference<br>[95% CI] | p-value |
| Site/school<sup>a</sup> |  |  |  |  |  |
| Bvumbwe | 19 | Ref. | 0.36 | Ref. | 0.20 |
| Ngowe | 24 | -39 [-93 to 470] |  | -50 [-89 to 123] |  |
| Maseya | 71 | -63 [-95 to 170] |  | -37 [-81 to 108] |  |
| Makhuwira | 65 | -79 [-97 to 70] |  | -70 [-91 to 1] |  |
| Season |  |  |  |  |  |
| Dry | 77 | Ref. | <0.001 | Ref. | <0.001 |
| Rainy | 102 | 4740 [2210 to 10040] |  | 5186 [2447 to 10870] |  |
| Age (per year increase) | 179 | -12 [-29 to 10] | 0.25 | -18 [-30 to -3] | 0.02 |
| Sex |  |  |  |  |  |
| Male | 92 | Ref. | 0.12 |  |  |
| Female | 87 | 150 [-20 to 710] |  |  |  |
| Hemoglobin |  |  |  |  |  |
| Not anemic | 137 | Ref. | 0.42 |  |  |
| Anemic<sup>b</sup> | 41 | 80 [-58 to 670] |  |  |  |
| Fever status<sup>c</sup> |  |  |  |  |  |
| Afebrile | 78 | Ref. | 0.13 |  |  |
| Febrile | 101 | 120 [-20 to 510] |  |  |  |
| Recent treatment<sup>d</sup> |  |  |  |  |  |
| Not treated | 168 | Ref. | 0.21 |  |  |
| Treated | 10 | 340 [-56 to 4240] |  |  |  |
| Slept under ITN<sup>e</sup> |  |  |  |  |  |
| No | 102 | Ref. | 0.30 |  |  |
| Yes | 73 | 90 [-43 to 510] |  |  |  |

\* Obtained in a survey weighted (weights calculated by school\*standard) linear regression with logarithm transformed gametocyte density as an outcome, and analyzing the variables shown in the table
\*\*Percent difference in gametocyte density from reference category is calculated as: $[\exp(\text{coeff}) - 1] \times 100$ .
ITN = insecticide treated net; OR = Odds Ratio; CI = Confidence Interval
<sup>a</sup> Univariate association for school, all bivariate include adjustment for school. <sup>b</sup> Hemoglobin <11.0g/dL; 1 missing observation <sup>c</sup> Measured temperature $\geq 37.5^{\circ}\text{C}$ at baseline visit or reported in the last two weeks <sup>d</sup> Reported in the last two weeks; 1 missing observations <sup>e</sup> 4 missing observations

### Gametocyte-containing infections missed by RDT screening

A key factor in the success of screen-and-treat interventions is the screening test sensitivity for detecting infections of interest, i.e., gametocyte-containing infections for transmission reduction. Sixteen percent of all students gametocyte-containing infections and 9% with high-density gametocyte-containing infections were RDT-negative and thus not treated. RDTs were less likely to detect gametocyte-containing infections at lower densities (Table 3). After adjusting for density, RDTs also missed gametocyte-containing infections more often in the lowest prevalence school (Bvumbwe) than other schools (OR for failure 6.6, 95% CI:1.9-24, p=0.004).

**Table 3:** Characteristics of gametocyte containing infections missed (not detected) by RDT screening at baseline.

| Predictor | N gametocyte positive | Weighted proportion <u>not</u> detected by RDT | Bivariate association*<br>OR [95% CI] | p-value | Multivariable association*<br>OR [95% CI] | p-value |
| --- | --- | --- | --- | --- | --- | --- |
| Site/school<sup>a</sup> |  |  |  |  |  |  |
| Bvumbwe | 19 | 0.40 | Ref. | 0.061 | Ref.<sup>g</sup> | **0.01** |
| Ngowe | 25 | 0.17 | 0.31 [0.07-1.4] |  | **0.21 [0.04-1.2]** |  |
| Maseya | 71 | 0.19 | 0.34 [0.10-1.2] |  | **0.25 [0.06-1.0]** |  |
| Makhuwira | 65 | 0.10 | 0.16 [0.04-0.60] |  | **0.09 [0.02-0.38]** |  |
| Season |  |  |  |  |  |  |
| Rainy | 103 | 0.11 | Ref. |  |  |  |
| Dry | 77 | 0.23 | **2.8 [1.2-6.8]** | **0.02** |  |  |
| Age (per year increase) | 180 | ---<sup>b</sup> | 1.1 [0.96-1.3] | 0.13 |  |  |
| Gametocyte density (log10) | 180 | ---<sup>c</sup> | **0.76 [0.64-0.90]** | **0.002** | **0.76 [0.64-0.90]** | **0.002** |
| Sex |  |  |  |  |  |  |
| Male | 92 | 0.13 | Ref. | 0.24 |  |  |
| Female | 88 | 0.20 | 1.6 [0.72-3.7] |  |  |  |
| Hemoglobin |  |  |  |  |  |  |
| Not anemic | 138 | 0.17 | Ref. | 0.83 |  |  |
| Anemic<sup>d</sup> | 41 | 0.13 | 0.89 [0.30-2.6] |  |  |  |
| Fever status<sup>e</sup> |  |  |  |  |  |  |
| Afebrile | 78 | 0.21 | Ref. | 0.39 |  |  |
| Febrile | 102 | 0.13 | 0.70 [0.31-1.6] |  |  |  |
| Recent treatment<sup>f</sup> |  |  |  |  |  |  |
| Not treated | 169 | 0.17 | Ref. | 0.39 |  |  |
| Treated | 10 | 0.07 | 0.40 [0.05-3.4] |  |  |  |
| Slept under ITN<sup>h</sup> |  |  |  |  |  |  |
| No | 102 | 0.13 | Ref. | 0.41 |  |  |
| Yes | 74 | 0.18 | 1.4 [0.60-3.5] |  |  |  |

\* Obtained in a survey weighted logistic regression of gametocyte-containing infections with the outcome of RDT negativity. All analyzed variables are
shown in the table.
ITN = insecticide treated net; OR = Odds Ratio; CI = Confidence Interval
<sup>a</sup> Univariate association for school, all other associations include school. Prevalence setting: Bvumbwe-low, seasonal; Ngowe-moderate, seasonal; Maseya-high; Makhuwira-high.
<sup>b</sup> Mean age of students with gametocyte containing infections not detected by RDT was 11.5 compared to 10.8 years for students with gametocyte containing infections detected by RDT.
<sup>c</sup> Among students with gametocyte-containing infections not detected by RDT, the mean log gametocyte density was -2.417 compared to a mean log gametocyte density of -0.825 among students with gametocyte-containing infections detected by RDT.
<sup>d</sup> Hemoglobin <11.0 g/dL; 1 missing observation
<sup>e</sup> Measured temperature $\geq 37.5^{\circ}\text{C}$ at baseline visit or reported in the last two weeks
<sup>f</sup> Reported in the last two weeks; 1 missing observation
<sup>g</sup> Adjusted OR for Bvumbwe compared to all other schools in the multivariable model is 6.6 [95%CI: 1.9-24], $p=0.004$

### Impact of screen-and-treat intervention on gametocyte prevalence and density in the school-based cohort

As hypothesized, treatment decreased gametocyte prevalence and density among students during follow-up. Comparing gametocyte prevalence among all students from baseline to six weeks after the intervention there was a reduction from 31.1% (95%CI: 27.6, 34.5) to 12.8% (95%: 9.5,16.1) in the rainy season and from 25.7 (95%CI: 20.9, 30.4) to 8.8% (95%: 5.4, 12.2) in the dry season. Geometric mean gametocyte density decreased from 0.50 gametocytes/microliter (95%CI: 0.09, 2.81) to 0.02 (95%: 0.002, 0.20) in the rainy season and 0.003 (95%CI: 0.001, 0.009) to 0.0001 (95%: 0.00001, 0.002) in the dry season.

The overall reduction occurred among treated students, while prevalence and density among untreated students remained constant or increased. The prevalence of gametocyte-containing infections among treated students decreased from 52% (95% CI: 45-58%) at baseline to 11% (95% CI: 7-14%) after two weeks, a 79% reduction (Figure 2A). Gametocyte prevalence remained low after six weeks [10%, 95% CI: 6-14%)]. Among untreated students (RDT-negative), the prevalence of gametocyte-containing infections remained unchanged from baseline [9% (95% CI: 5-14%)] to six weeks after the intervention [12% (95% CI: 7-17%)].

**Figure 2:**
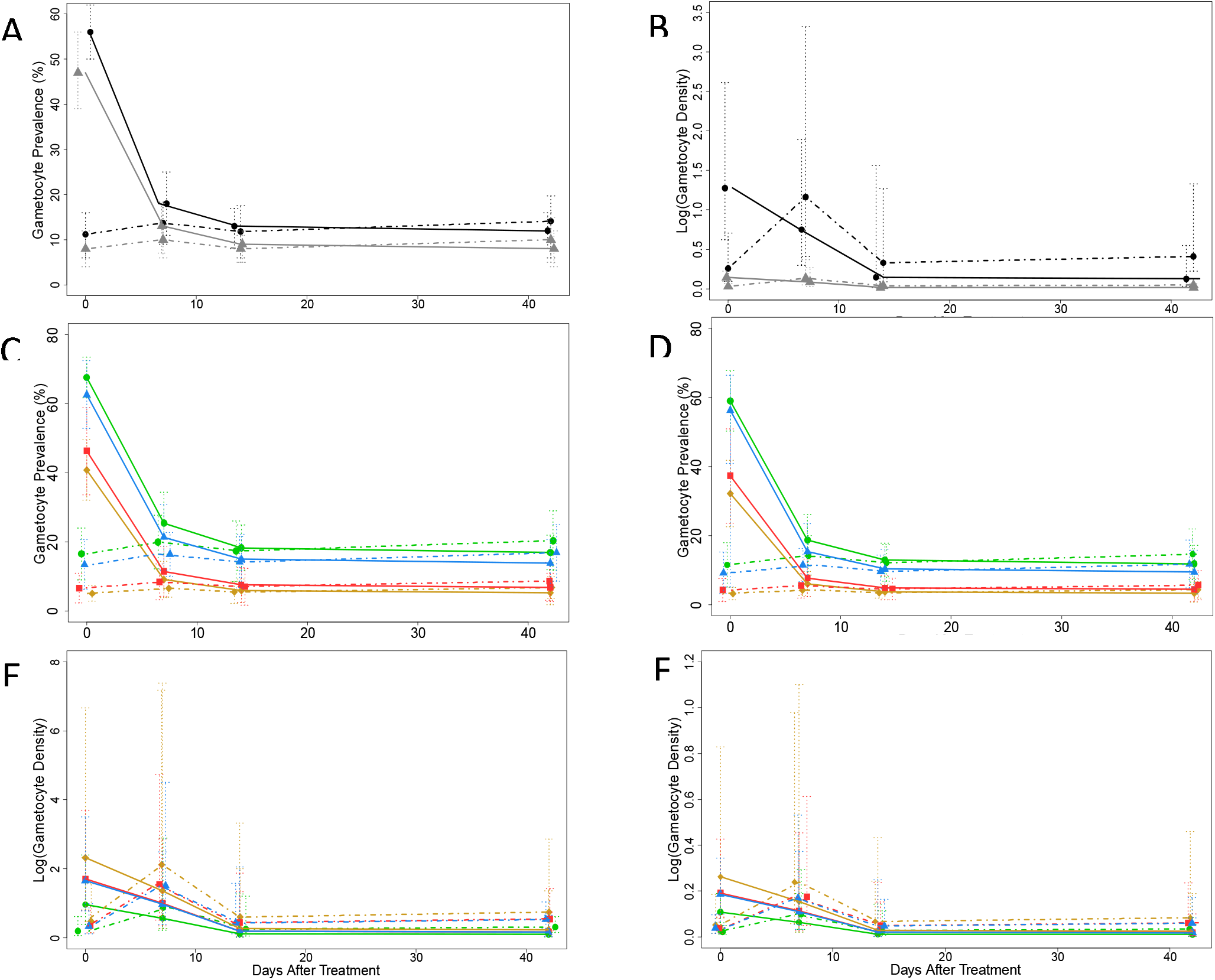
Impact of treating RDT+ students on the proportion and density of gametocytes in the school-based cohort over time. Proportion of students with gametocytes in all schools combined (A) and by school in the rainy (C) and the dry (D) season. Mean gametocyte density (logarithm transformed) among students in all schools (B) and by school in the rainy (E) and the dry (F) season. Solid lines represent students who received treatment; Dashed lines represent students who did not receive treatment. Color designates season in A and B rainy (black and dry (gray). Color designates school in C through F Bvumbwe (yellow), Ngowe (red), Maseya (blue), Makhuwira (green). Both estimates were obtained in random effects longitudinal analysis (full models in Supplemental Tables S3 and S4).

Gametocyte density followed the same trend, with 90% reduction in the geometric mean of gametocyte density after treatment versus 59% increase in untreated children (Figure 2B). There were 32 students with high-density gametocyte-containing infections at baseline: 28 in RDT-positive students and four in RDT-negative students. Two weeks after screening-and-treatment, only one high-density gametocyte infection was detected, which was in an untreated student. Six weeks after treatment there were four high-density gametocyte-containing infections, two in students that were not treated and two in students that received treatment, corresponding to a 90% reduction in the prevalence of high-density gametocyte infections. Results were similar across schools, a proxy for transmission settings (Figures 2C-F).

### Gametocytemia in the school-based cohort compared to school-age children in the community

The proportion of infections containing gametocytes in the school-based cohort was similar to that in school-age children in the community (25% vs. 27%, p=0.62), supporting the generalizability of the intervention results to all school-age children in the community. The geometric mean density was higher in school-age children in the community (0.99 gametocytes/µl [95% CI:0.71-1.38]) than in the school (0.34 [95% CI:0.19-0.60], p=0.03). However, the proportion of infections containing ≥10 gametocytes/µl among those with gametocytes was similar (20% in the community vs. 18% in schools, p=0.51).

### Predicted reduction in the community gametocyte population following treatment in schools

The screen-and-treat intervention reduced gametocytemia in our school-based cohorts. Furthermore, school-age children substantially contributed to the population of gametocyte-carriers in the surrounding communities; 46% of all gametocyte-containing infections and high-density gametocyte-containing infections were in school-age children, who comprised only 35% of the population (Supplemental Results and Figure S2). If the screen-and-treat intervention were extended to all school attendees, we estimate it could result in at least six weeks of reductions in the community prevalence of gametocyte-containing infections as large as 26% in the rainy and 34% in the dry season (Figure 3A and 3B). The total gametocyte burden (sum of gametocyte densities) in the community would be reduced by 33% in the rainy season and 25% in the dry season (Figure 3C and 3D). The number of infections containing ≥10 gametocytes/µl would be reduced by 44% and 55% in the rainy and dry seasons, respectively (Figure 3E and 3F).

**Figure 3:**
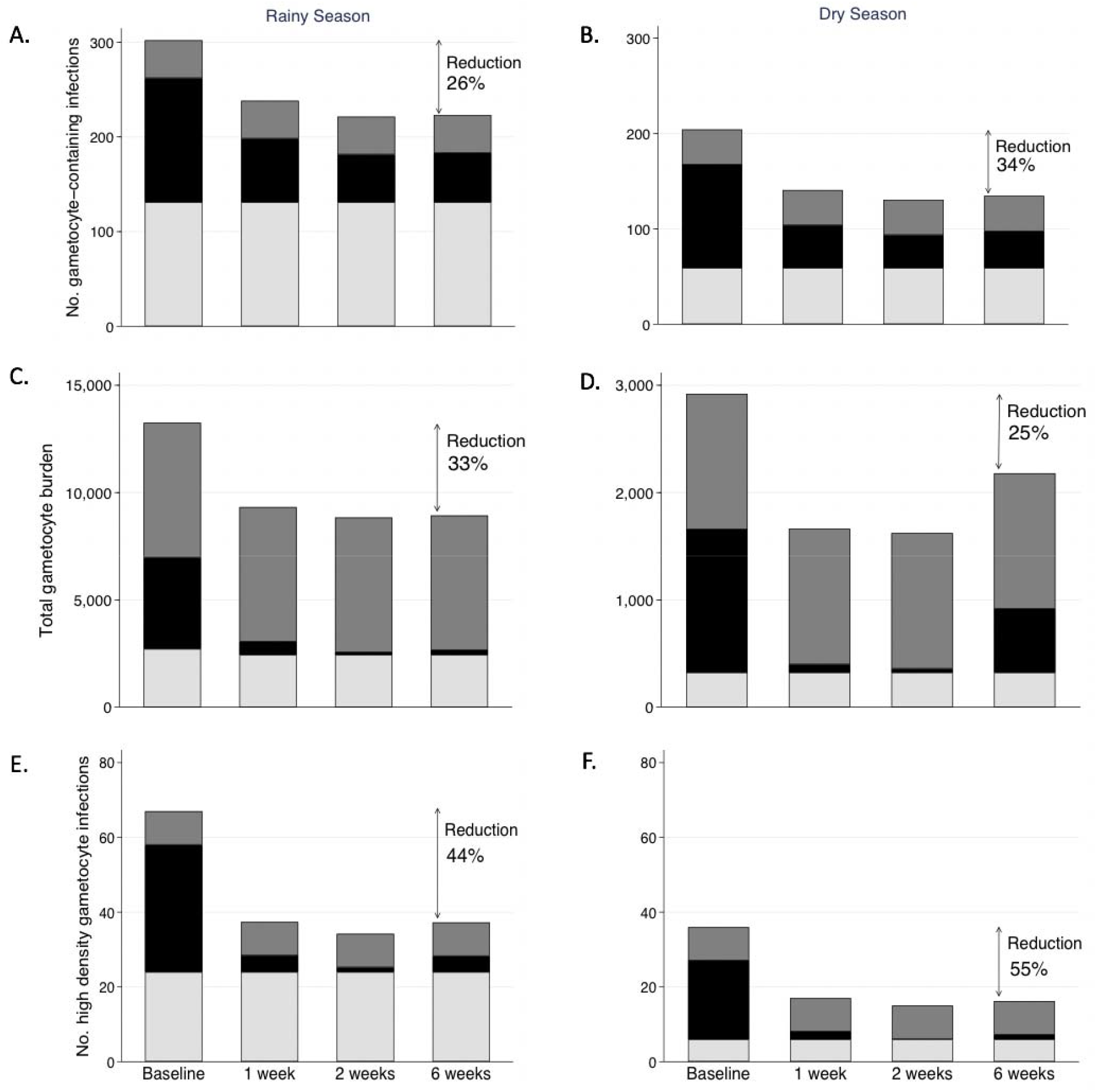
Predicted impact of school-based treatment of RDT positive students on the population of gametocytes in the surrounding community. Following school-based screening-and-treatment at baseline, the estimated impact on gametocyte prevalence (A in rainy and B in dry seasons), total gametocyte burden (C in rainy and D in dry seasons), and number of infections containing ≥10 gametocytes/µl (E in rainy and F in dry seasons) in the communities surrounding the schools are predicted at one, two, and six weeks after the intervention. Color indicates the proportion of the gametocyte measure by age group: school-age children (6-15y) – black; younger children (6-71m) – light grey; adults (>15y) – dark grey. Total gametocyte burden is the sum of gametocyte densities in individuals in each age group. These calculations assume treatment is not provided to young children, adults, or school-age children who test negative by RDT when the intervention is implemented. Reduction is calculated as the proportional difference between the baseline and six-weeks post intervention.

## Discussion

Additional malaria control interventions are needed in areas like Malawi where current measures have not yielded substantial reduction in *P. falciparum* transmission. Our results support the use of school-based screen-and-treat as an intervention that may not only improve the health of schoolchildren, but also decrease transmission in the community. Almost half of high-density gametocyte-containing infections in the community were in school-age children; school-based treatment nearly eliminated this group’s contribution to transmission for up to six weeks. We also showed that RDTs detected most high-density gametocyte-containing infections, and that treatment of sub-clinical infections based on RDT results reduced gametocyte prevalence and density. Treating only children with positive RDTs could reduce the community-level gametocyte burden by a third. This reduction in potentially infectious reservoirs could help interrupt persistent malaria transmission.

The high prevalence of gametocyte-containing infections in school-age children in this study is consistent with prior work by our group and others.^8,11,16^ However, to our knowledge, this is the first study to simultaneously determine gametocyte burden in schools and surrounding communities to estimate the potential impact of school-based treatment on transmission. In Uganda, school-based intermittent preventive treatment was associated with reduced parasite prevalence in the surrounding community in a large cluster randomized trial.^23^ Although the community-level effect was statistically significant, the magnitude was limited by low intervention coverage. Because this intervention was conducted as a clinical trial, parents had to come to the school to consent for each student, creating logistical barriers. Thus, further estimates of the full potential of this intervention are needed.

While our results lend further support the potential contribution of school-based preventive treatment to transmission reduction, there are limitations to our analyses. We did not use RDTs to detect infections in the communities surrounding schools and had to extrapolate from the relationships of PCR- and RDT-detected infections in the school-based cohort to estimate the number of RDT-positive children who would attend school from the community. However, the distribution of gametocyte prevalence in the school-based and community-based surveys was comparable, supporting the estimate. Furthermore, we used sensitive molecular methods to detect and quantify gametocytes but did not directly measure infectiousness using feeding assays. However, our data still likely represent the relative contribution of different age groups to transmission because we evaluated not only the prevalence of gametocytes but the density and further evaluate high-density gametocyte containing infections, which are the most likely to lead to mosquito infection.^24^ Furthermore, prior studies have shown that school-age children and younger children are more infectious to mosquitos than adults^12^ and school-age children are more likely than younger children to be bitten by competent vectors.^16^ Lastly, our estimates of the impact of the intervention in the community assume universal coverage in students. In Malawi, school-based deworming programs routinely report coverage of >90% of school-age children. ^25^ Thus high levels of coverage are possible.

Our results likely underestimate the overall effect of school-based treatment for three reasons. First, the impact of clearing gametocyte-containing infections in school-age children is expected to be amplified because competent vectors probably feed more often on school-age children and adults who have larger body surface than younger children^15,26^ and use bed nets less frequently.^14–16,26^ Second, in our community-level predictions, we conservatively assumed that gametocyte-containing infections in non-treated age groups would remain constant. However, the decreased pool of infectious school-age children should also lead to fewer new infections and fewer gametocyte-containing infections in younger children and adults. Third, our results are based on a single treatment intervention; the impact of repeated school-based treatment is likely to be even larger. In clinical trials that have evaluated the impact of school-based malaria preventive treatment on health outcomes, the most successful interventions were repeated either monthly during the high transmission season or quarterly throughout the year.^20^ Reinfection rates, seasonal variation in transmission, and duration of the prophylactic effect of the treatment drugs should be used to guide the frequency of intervention required.

Currently, schools are platforms for successful deworming campaigns and feeding programs, and a growing number of interventions target this age group, including vitamin supplementation and adolescent vaccinations. The vast majority of children in sub-Saharan Africa attend school, making schools a logistically feasible platform for targeted interventions. Schools are particularly important to reaching this age group, since children rarely have routine contact with the health care system after completing childhood immunizations. Adding malaria preventive treatment to other school-based interventions in an integrated school health package should improve the cost:benefit ratio, promote sustainability, and improve health care access to an underserved population.

One concern is that risk of infection and disease could increase once students leave school or if the intervention ceased. A prior study of chemoprophylaxis in younger children demonstrated a transient increase in clinical infection when the intervention was discontinued.^27^ However, this “rebound effect” was not observed in all chemoprophylaxis trials, nor has it been found after intermittent preventive treatment of infants.^28–30^ In highly endemic settings where school-based treatment interventions are most needed, schoolchildren have acquired partial immunity and often have sub-clinical infections. Continued exposure should maintain some naturally acquired immunity. Another concern about the widespread use of preventive treatment is the potential for drug resistance. While this concern is important, it should be weighed against the direct student health and potential indirect community benefits when evaluating the approach.^20^

Our results join previous studies in supporting the need for implementation studies to measure the indirect impact of school-based treatment on community-level malaria transmission. Although RDTs will fail to detect some low-density infections, such as those observed in the dry season and in our lowest prevalence school, a screen-and-treat approach may substantially reduce the population of gametocytes in the community. Alternatively, providing preventive treatment to all students as intermittent preventive treatment, may have even greater impact. While higher density gametocyte-containing infections are more likely infectious, low density infections contribute to population-level transmission dynamics because they are common.^24^

In summary, these results demonstrate the potential for school-based malaria treatment to substantially decrease *P. falciparum* gametocyte carriage and thus transmission in the surrounding community. In combination with disease- and vector-control interventions, this transmission-focused strategy may be critical to reduce the burden of malaria in areas where this disease has remained entrenched despite current control measures.

## Methods

### Study sites and design

School-based cohort studies and household-based cross-sectional surveys were conducted at the end of the rainy season (April-May) and during the dry season (September-October) of 2015. Two sites with consistently high (>40%) parasite prevalence in school-age children – Maseya and Makhuwira – and two with lower, seasonally varied transmission (>two-fold seasonal prevalence difference) – Bvumbwe and Ngowe – were selected from 30 previously studied sites in southern Malawi.^6^ Long-lasting insecticide treated nets were distributed through a national campaign in 2012. Rapid diagnostic tests (RDTs) and treatment with artemether-lumefantrine were generally available in local government-operated health facilities.

#### School-based cohorts

Students present on a sampling day were assigned numbers. Fifteen students per grade-level were sampled using a random number generator. Students were excluded if they: had no parent/guardian available to provide consent, would not attend school throughout the 6-week study, or had a known artemether-lumefantrine allergy. Students enrolled in the rainy season cohort were excluded from the dry season cohort.

Enrolled students were interviewed at baseline, one-, two-, and six-week visits about bed net use the night prior, current or recent illness, and antimalarial treatment. At baseline, a finger-prick blood sample was obtained for detection of *P. falciparum* by RDTs and hemoglobin was measured by portable photometer (Hemocue, Angelholm, Sweden). RDT-positive students received weight-based treatment with artemether-lumefantrine (Novartis Pharma AG or Ajanta Pharma Ltd). At baseline and all follow-up visits, finger-prick blood was obtained for molecular detection of any-stage parasite and gametocytes. At the final visit, parents were interviewed and health passports (individual portable medical records) reviewed to identify intercurrent fever or malaria treatment.

#### Household-based cross-sectional surveys

Concurrently cross-sectional surveys were conducted in 80 households in each school catchment area, including a cluster of 30 from ongoing surveillance studies (Details in ^6^) and 50 households newly selected based on closest Euclidian distance to the school. Households were visited within two weeks of the school cohort baseline visit. Questionnaires based on the Malaria Indicator Survey were administered and blood samples obtained for polymerase chain reaction (PCR) testing, as previously described.^6,14^ RDTs were not performed and no treatment was provided. School attendance in the last four weeks was asked for all participants ≥ five years. Few five-year-olds attended school, thus school-age children were defined as those six to 15 years old. Younger children were defined as 6-71-months-old, and adults as over 15 years old.

### Ethical considerations

Written informed consent and assent were obtained from adults and children, as appropriate. The University of Malawi College of Medicine Research and Ethics Committee and the Institutional Review Board of the University of Maryland Baltimore approved this study.

### *Plasmodium* infection detection methods

At the school-based study baseline visit a histidine-rich protein 2-based RDT (Paracheck® Orchid Biomedical Systems, Goa, India or SD Bioline, Standard Diagnostics Inc., Suwon City, Republic of Korea) was used per manufacturer instructions. For all blood specimens, real-time PCR was performed to detect *P. falciparum* lactate dehydrogenase (*Pf*LDH) DNA, as described previously.^11^ Plates were run in duplicate. If *Pf*LDH positive in either run, quantitative reverse transcription PCR was performed on whole blood preserved in RNAprotect Cell Reagent (Qiagen Inc. Valencia, CA, USA) to measure expression of the mature gametocyte marker *Pfs25*.^31^

### Infection and treatment definitions

*P. falciparum* infection was defined as positive *Pf*LDH PCR results on at least one run. Participants without *Pf*LDH detected by PCR were considered gametocyte negative. RDT-positive students with negative PCRs were considered negative for active infection, but were included in the treatment group for analysis because they had received artemether-lumefantrine. Other malaria treatment during follow-up was defined as any treatment with an effective antimalarial reported during student follow-up interviews, during the parent interview, or in the student’s health passport. High-density gametocyte infections were those with ≥ 10 gametocytes/microliter (µl). Lower densities of gametocytes are thought to contribute less to transmission [25].

### Statistical analysis

All analyses of the school-based population accounted for survey weights and school sampling strata (Supplementary Materials). Contribution to transmission was evaluated four ways: 1) gametocyte prevalence defined as the proportion of participants with any gametocytes; 2) gametocyte density; 3) total gametocyte burden calculated as the sum of participant gametocyte densities; and 4) prevalence of high-density gametocyte infections.

For school-based cohorts at baseline, logistic regression models were used to assess determinants of gametocyte prevalence and linear models (log10-transformed) were used to assess determinants of gametocyte density y among infections with > 0 gametocytes. We used logistic regression among students with gametocyte-containing infections to identify characteristics of students in whom RDTs failed to detect gametocyte-containing infections at baseline. Gametocyte density was included as a continuous variable after assessing linearity of the relationship with odds of RDT negativity. Because of baseline differences in transmission pattern and demographics, all analyses were adjusted for school. In all models, selection of relevant predictors was based on Wald tests through best subset regression. Effect in linear models was reported as percent difference in absolute gametocyte density and was calculated as [exp(coeff)-1] x 100.

Differences in the prevalence of gametocytemia and gametocyte density over time between treated (RDT positive) and untreated students (RDT negative) were assessed in nested random intercept longitudinal models by testing an interaction term between treatment and indicator variables for visit. Based on these longitudinal models, predictions of gametocyte prevalence and density were obtained, marginal on random intercepts, and stratified by school, season, and treatment status (baseline RDT positive or negative), with other modeled predictors assumed constant.

To assess the contribution of school-age children to the overall community population of gametocytes, we compared the distribution of gametocyte-containing infections, high density gametocyte infection, and geometric mean gametocyte density through chi-squared tests and linear regression: i) across age-groups, and ii) between students in the school-cohort and school-age children in the community. This analysis was unweighted because sampling of community surveys was not stratified.

### Predictive modeling

To evaluate the potential indirect impact of school-based preventive treatment on the community, we estimated the expected reduction in the number of gametocyte-containing infections, gametocyte burden, and the number of high density gametocyte-containing infections in the communities surrounding schools if the screen-and-treat intervention had included all students. Briefly, we first calculated the number of children from the community likely to be in school from the age- and gender-weighted proportion of school-age children that reported attending school in the past four weeks. To estimate the expected reduction in the numbers of gametocyte-containing infections and high-density gametocyte-containing infections in the communities surrounding schools, we used mixture of distributions and conditional probabilities. To estimate the impact of the intervention on the overall gametocyte burden, we first fit overdispersed Poisson models to the school-based cohort with gametocyte density and RDT positivity results at baseline, then predicted the overall gametocyte density changes following the intervention for the hypothetical population of school-going children from the community who would have had a malaria infection and been treated in the screen-and-treat intervention (RDT-positive at baseline) at each follow-up time point. We assumed the population prevalence of gametocyte-containing infections in adults and younger children would remain constant during the intervention period because they would not receive the intervention.

Analyses were conducted in Stata/SE version 15.1 (StataCorp, College Station, TX, USA) and R v3.5 (R Foundation for Statistical Computing, Vienna, Austria).^32^ Weighted analyses used the survey module of Stata. Significance was set at alpha = 0.05. Further details of the predictive modeling and statistical analysis are provided in the Supplemental Information.

## Data Availability

Data will be publicly available in association with the Malawi International Center of Excellence in Malaria Research (U19AI089683).

## Acknowledgements

We appreciate valuable input on the analysis and crafting of the manuscript from Drs. Kristin Stafford, Sania Amr, and Andrea Buchwald. We thank Heidi Fancher and the administrative team at the Malaria Alert Center for their support in executing the study. We are grateful for the hard work and dedication of the field team, study nurses, teachers, school administrators who made the study possible. Most importantly, we thank the participants for their commitment and patience.

## Contributors

Conceived of the study concept and design: LMC, CV, MLW, MKL. Data collection and management: LMC, JEC, MC, ANg, AB, TET, DPM, MKL. Laboratory analysis: ANy, KBS. Performed data analysis: LMC, CV. Wrote the first draft of the manuscript: LMC, CV, MLK. Agree with manuscript results and conclusions: LMC, CV, JEC, ANy, MC, ANg, AB, KBS, MLW, TET, DPM, MKL

