## Supplemental Information for "School-based screening and treatment may reduce *P. falciparum* transmission"

**Contents:**

**Additional Methods**

Text S1**:** Study design and Data collection

Text S2: Laboratory analysis

Text S3: Statistical analysis

**Figures and Tables**

Figure S1: Enrollment Flowchart

Table S1: STROBE Checklist

Table S2: Baseline prevalence and predictors of high density gametocyte infections (≥10 gametocytes/microliter) among infections containing gametocytes at baseline

Table S3: Survey weighted longitudinal mixed-effects logistic regression model of gametocyte prevalence with random effects for strata (school and grade) and student within strata

Table S4: Survey weighted longitudinal mixed-effects linear regression model of gametocyte density (gametocytes per microliter) with random effects for student

Figure S2: Age distribution of gametocyte-containing infections in communities surrounding schools

**Results**

Text S4: Contribution of school-age children to the population of gametocytes in the communities surrounding schools

**Additional methods**

**Study design and Data collection**

Prior to initiating the study, sensitization meetings were held with community leaders, school leaders, and district and local government officials. Local community and school leaders provided permission to survey each community and school.

Sample size was based on reduction of the prevalence of infection in students after the screen-and-treat intervention. Based on prior surveillance data in the communities surrounding the intervention schools, we estimated there would be overall 27% prevalence of infection detected by PCR in school-age children. By sampling 320 (10 students/grade/school in each season), we would have 80% power to detect a 40% decrease in the prevalence of infection after the screen-and-treat intervention. We also estimated that, using prevalence by microscopy as a proxy for RDT positivity, we would identify 64 RDT positive individuals (20% microscopy positive) and 22 individuals who are RDT negative, but PCR positive (7% sub-RDT infections). Using qRTPCR, we have detected gametocytes in ~50% of children who are PCR positive, so we anticipated detecting 43 infections containing gametocytes. To account for loss to follow-up, we sampled 15 students/grade in each school in the rainy and the dry season cohorts.

Questionnaires were administered in Chichewa, the local language, on android-based tablets using OpenDataKit ([http://opendatakit.org](http://opendatakit.org/)) and managed using electronic data capture.

**Laboratory analysis**

Gametocyte detection and quantification: RNA was extracted using RNeasy Plus Mini-Kits® (Qiagen Inc.,Valencia, CA) and treated with RNase-free DNase Sets®(Qiagen Inc., Valencia, CA) to eliminate parasite gDNA. Reverse transcription and PCR were performed using TaqMan RNA-to-Ct 1-Step Kits (Applied Biosystems, Foster City, CA). Twenty microliter reactions were performed in duplicate each including 2µl sample RNA. Samples were considered positive if either replicate was positive. Gametocyte density was quantified using a standard curve of gametocytes induced from Pf.2004 TdT *P. falciparum* (provided by the lab of Dr. Matthias Marti). The quantity of gametocytes in the standard curve was microscopically determined by three independent readers, then diluted using uninfected red blood cells to create a stock standard with a concentration of 50,000 gametocytes/μL. Standard stock was diluted to 200 gametocytes/μL for the high-density control and serially diluted (5-fold) to 5.12x10^-4^ for development of a standard curve that was run on each plate. Plates were repeated if sensitivity of 2.56 gametocytes/mL was not achieved.

**Statistical analysis**

**School-based:**

Survey weights were calculated for each stratum, defined as each grade within each school [N_totalstudents_/n_sampledstudents_] (Levy 2008). For example, if the total number of students in standard 1 in school #1 is 150 and we sampled 15 students from that standard then the weight for that strata is 150/15=10. To allow estimations of gametocyte density, strata were collapsed to ensure one observation per strata. No more than two strata were combined. Final analyses included 26 strata. In longitudinal analysis, survey weights were normalized.

In longitudinal analysis, changes in trends over time across schools and between seasons could not be assessed because models including interactions did not converge. However, to explore whether the assumption of a parallel effect of treatment on outcomes over time was consistent across schools and seasons (i.e., if model without interaction yield valid inferences), we investigated observed proportions and means of each subgroup over time. Those observed values were consistent with the prediction obtained by models and presented in Figure 2.

Logistic longitudinal mixed models for prevalence of gametocytemia included a random intercept for student nested in a random intercept for school strata. However, to keep models parsimonious, when constructing longitudinal models for gametocyte density, the random intercept for sampling strata was removed from the model and final models only included a random intercept for child because the variance of the random intercept for strata was nearly zero (0.0007). Moreover, although we could not use restricted maximum likelihood (REML) to compare survey longitudinal models with and without the random intercept for school, the p-value of REML was 0.95 when comparing of unweighted model with a random intercept for child only and the unweighted model with a random intercept for child nested in sampling strata. While the final fitted models did not include sample weights, they had results very similar to weighted models, and thus, these comparisons were considered valid. Finally, predictions of gametocyte prevalence and density were obtained in longitudinal models. In these models, the adjusted predictions at representative values that were plotted in Figure 2 corresponded to strata specific predictions of fixed effects for all predictors included in models, marginal on random intercepts. Additionally, we estimated in these models the average marginal effects that were reported in the text for overall estimates marginal across schools and seasons. The average marginal effects average the predicted outcome of a subject assuming that the subject changes the values of one predictor, given all other predictors remain constant.

Survey weighted descriptive statistics including means and proportions, and regression analysis were calculated using the svy command in STATA. In longitudinal analysis, survey weights were normalized.

**Household-based:**

The age distribution of infections containing gametocytes was calculated as the proportion of gametocyte containing infections in each age group (young children 6-71m, school-age children 6-15yo, and adults >15yo). Proportions were compared using the chi-squared test. The same approach was used to compare infections containing >10 gametocytes/microliter by age group.

**Prediction of impact of school-based treatment on gametocytemia in school-aged children from the community**

Comparisons of the observed gametocyte carriage (proportion and burden [sum of participant gametocyte densities]) in the community served by each school to what we expected the gametocyte carriage would be 1, 2, and 6 weeks after a school-based malaria treatment intervention in the high and low transmission seasons were carried out based on observed data and estimated proportions obtained in the school-based and in the community-based surveys.

For all calculations, the number of school-aged children in the community and the estimated proportion of school attendance of those children were obtained in the community-based cross sectional survey. School attendance was evaluated by asking parents during the community-based survey whether children 5-15 years old had attended school in the last four weeks. Detailed calculations are presented below. In blue are quantities used for these calculations obtained in the community-based survey and in red are quantities obtained in the school-based survey. In purple are estimates combining data from the community and the school.

***Impact on gametocyte prevalence in communities***

This analysis required estimating two quantities: a) the number of gametocyte carriers in the rainy and dry seasons (*k = 1 for rainy and 2 for dry*), in the absence of an intervention at baseline (*t =* 0), in the school-aged population of each community (sac) served by each *i* of the four schools (*i* = 1….4), i.e.,
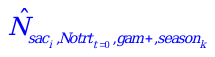
; b) the number of gametocyte carriers in a given season at one, two, and six weeks after treatment (*t=j* for *j*=1, 2, and 6 weeks) in the school-aged children of the communities served by each school if RDT positive children would have received treatment in the school at baseline (*t =* 0), i.e.,
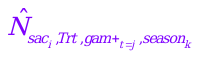
.

First, to estimate
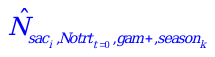
, we assumed that it was binomially distributed and was constant for every *t=j* , i.e., it was the same 1, 2, and 6 weeks after treatment.

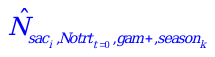
~
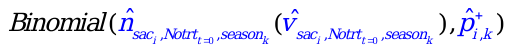
 (1)

In this distribution, the quantity
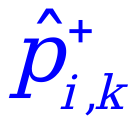
represents the proportion of gametocyte positive school-aged children in the community of school *i* in a given season *k* or
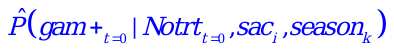
at baseline. That quantity was estimated in the cross-sectional community-based survey (and thus, is in blue) at one point in time and assumed to be constant for *t* = 1,2 and 6 weeks. The quantity
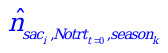
was the number of school-aged children that would have attended school and thus would be candidate to the intervention in each of the four communities. This last quantity was obtained by combining the proportion of school-aged children that attended school in each of the four communities (range 0.82 to 0.99) with the observed number of school-aged children in each of the four communities in each season (range 101 to 157). Both of these last quantities were measured in the cross-sectional community-based survey. This
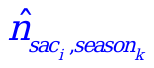
was also assumed to be binomially distributed, constant over time, and was a function of two quantities:

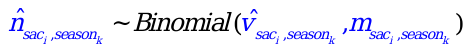
 (2)

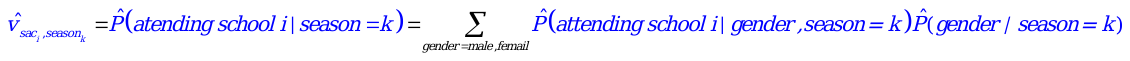
. In other words,
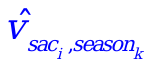
represents the proportion of school-aged children attending schools in each community that was estimated in the cross-sectional community-based survey. That proportion was found to vary by gender and, thus, was weighted over gender. The quantity
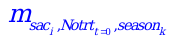
was the observed population of school-aged children in each community at each season. Therefore,
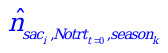
was estimated through the product
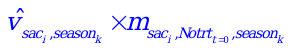
,
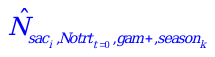
was estimated through the product
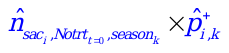
.

After obtaining
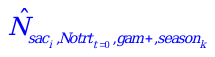
, we estimated the number of gametocyte carriers in a given season and post-treatment week (*t=j,* for *j*=1, 2, 6 weeks) in the school-aged children of the communities served by each school if RDT positive children would have received treatment in the school, i.e.,
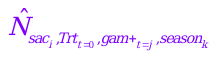
, also binomial but varying over time.

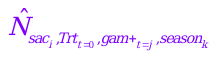
~
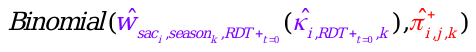
 (4)

The estimated proportion
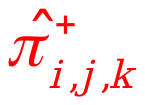
 was the proportion of gametocyte-positive children among treated children (RDT-positive at baseline) in each school and season at each visit (
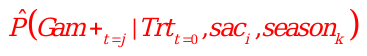
). To obtain this proportion, we used the estimated predictions from the logistic random intercept model of the school-based longitudinal study (Figure 3). Notice that Trt*_t_*_=0_ is equivalent in this case to RDT-positive. The quantity
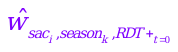
 corresponds to the population of school-aged children who would have undergone the intervention in schools because they were RDT-positive at baseline. Since we did not do RDTs in the community-based surveys, we estimated this population by assuming it was binomially distributed:

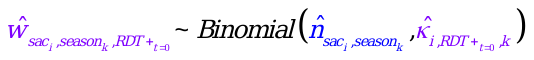
(5)

Therefore, estimation of
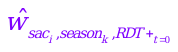
 was a function of: a) the school-aged population of a given school and season after the intervention (
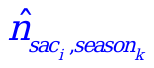
) that could be assumed to be equivalent to the population in the absence of the intervention (
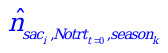
) that we obtained above; b) the proportion of children who would have received treatment, i.e., the probability of presenting a positive RDT result at baseline in each school and season. (
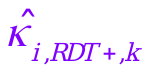
 or
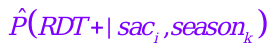
). Because the probability of a RDT-positive result is generally a function of PCR positivity (0 for negative and 1 for positive) for malaria parasites that was available in the community-based survey, we estimated the probability of an RDT-positive result in school-aged children in the community based on a mixture of community-based and school-based survey data as:

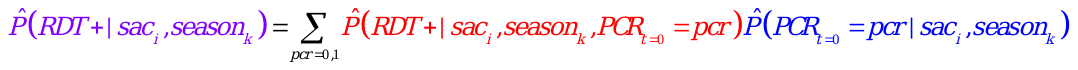
(6)

Where the conditional probability of RDT positivity on PCR positivity is plugged in from the school-based survey estimate and the conditional probability of PCR positivity is plugged from the community based survey. Estimating the probability of RDT positivity in this way was done because the probability of having a positive RDT in subjects with positive and negative are heterogeneous.

***Impact on total burden (sum of participants gametocyte densities) of gametocytes in communities***

This analysis required estimating the sum of participant density of gametocytes based on the following:

a) Sum of gametocyte densities among school-age children from the communities surrounding each of the *i* (*i* = 1…4) schools who were expected to attend schools in absence of treatment, in the rainy and dry seasons (*k = 1 for rainy and 2 for dry*), at baseline (*t=0*) and at the *t* subsequent visits (*t* = 1, 2 and 6 weeks)
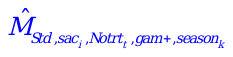

b) Sum of gametocyte densities among school-age children from the communities surrounding each of the *i* (*i* = 1…4) schools who were expected to attend schools that would have been treated had they had a RDT+, in the rainy and dry seasons (*k = 1 for rainy and 2 for dry*)
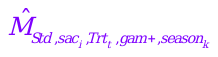
at the subsequent visits after treatment (*t* = 1, 2, and 6 weeks)
c) Sum of gametocyte densities among pre-school children in the community-based study from the communities surrounding school *sac* served by each of the *i* four schools (*i* = 1…4) in absence of treatment, in the rainy and dry seasons (*k = 1 for rainy and 2 for dry*)
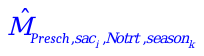
. Since pre-school children were not treated, this quantity was assumed to be constant over time.
d) Sum of gametocyte densities among adults in the community-based study from the communities surrounding school *sac* served by each of the *i* four schools (*i* = 1…4) in absence of treatment, in the rainy and dry seasons (*k = 1 for rainy and 2 for dry*)
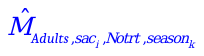
. Since adults children were not treated, this quantity was assumed to be constant over time.

For consistency, we obtained the sum of the gametocyte densities of all subgroups and at all visits using the same approach to generate the predictions. Because treatment influenced both the number of gametocyte-containing infections and the density of gametocyte-containing infections, determining the number of participants density estimates apply to is required. Thus, we estimated the density-based on predictions of zero inflated Poisson models fit with gametocyte density (including 0 density) in the original scale as outcomes. An ad hoc fit of these models was evaluated by comparing predicted and observed totals for each subgroup (in the school-based cohort, when analyzing school age-children, and in the community-based survey, when analyzing adults and pre-school children). Before we carried out analysis to obtain the quantities above, we deleted two influential extreme outliers who had gametocyte densities above the 99.9^th^ percentile of the total gametocyte distribution considering all visits of the school-based cohort and community-based surveys. In school-age children ((a) and (b) above), we used predictions based on hypothetical populations estimated as described below.

To obtain
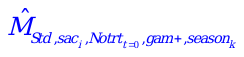
 we first fit a zero inflated Poisson model including all school-age children from the community-based survey, regardless of their school attendance. In these models, the gametocyte-density was the outcome, and season and school were predictors. Next we obtained the hypothetical population of school-age children from the community that was expected to attend school and were gametocyte positive and negative in the community of each school and season. To do that, we created a dataset for this population based on each
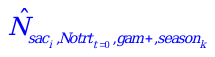
and
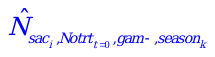
(described above), i.e., the season- and school-specific number of gametocyte positive and negative children expected to attend schools in the communities. Using the new population, we obtained the predicted gametocyte density in each child of this population and added it.

To obtain
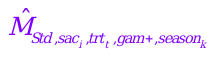
, we fit a zero inflated Poisson model at each visit *t* separately for the school-based cohort. In these models, gametocyte density was the outcome and treatment (i.e., RDT positivity), season, and school were predictors. As we did for the baseline estimates described above, we then obtained the hypothetical population of school-age children from the community who were expected to have attended the corresponding school, have been treated and untreated at baseline (RDT-positive and negative at baseline), and were gametocyte positive and negative in each season. To do that, we created a dataset for this population based on each
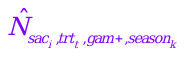
and
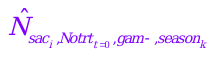
estimated above. Finally using the fitted models, we obtained the expected gametocyte density for this hypothetical (“new”) populations and added all children’s densities.

For young children and adults, respectively,
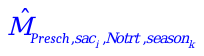
and
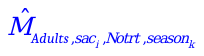
were estimated by fitting zero inflated Poisson models with the community-based data and predicting the gametocyte density using the same data.

***Impact on prevalence of high density gametocyte-containing infections in communities***

We used the same methods described above for the impact on gametocyte prevalence to estimate but in this case
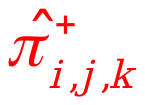
 represented the proportion of high density ($\geq$10 gametocytes/microliter) gametocyte-positive children among treated children (RDT-positive at baseline) in each school and season at each visit

(
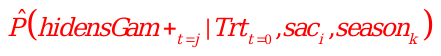
). Due to the smaller sample size for the high-density gametocyte-containing infection outcome, regression models did not converge. Thus, to estimate n
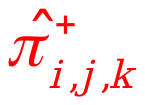
 we used the raw proportions from the school-based data by school, season, and baseline RDT result.

**Figure S1: Enrollment and follow-up of school-based cohorts**

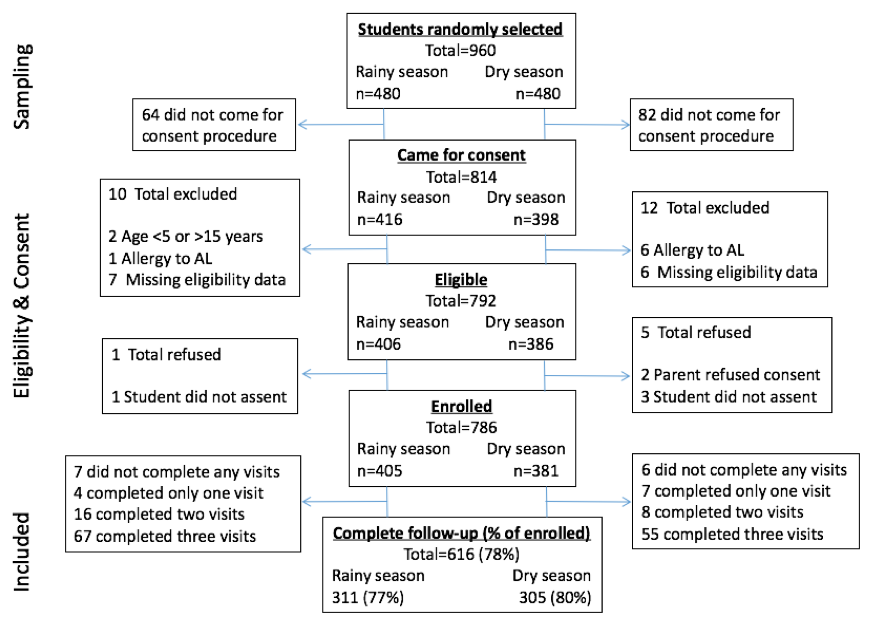

| Table S1: STROBE Checklist | | |  |
| --- | --- | --- | --- |
| **Section/topic** | Item No | Checklist item | Reported on page # |
| **Title and abstract** | 1 | (*a*) Indicate the study’s design with a commonly used term in the title or the abstract | 2 |
|  |  | (*b*) Provide in the abstract an informative and balanced summary of what was done and what was found | 2 |
| Introduction | | |  |
| Background/rationale | 2 | Explain the scientific background and rationale for the investigation being reported | 3-4 |
| Objectives | 3 | State specific objectives, including any prespecified hypotheses | 4 |
| Methods | | |  |
| Study design | 4 | Present key elements of study design early in the paper | 18-19 |
| Setting | 5 | Describe the setting, locations, and relevant dates, including periods of recruitment, exposure, follow-up, and data collection | 18-19 |
| Participants | 6 | (*a*) Give the eligibility criteria, and the sources and methods of selection of participants. Describe methods of follow-up | 18-19 |
|  |  | (*b*) For matched studies, give matching criteria and number of exposed and unexposed |  |
| Variables | 7 | Clearly define all outcomes, exposures, predictors, potential confounders, and effect modifiers. Give diagnostic criteria, if applicable | 19-23 |
| Data sources/ measurement | 8* | For each variable of interest, give sources of data and details of methods of assessment (measurement). Describe comparability of assessment methods if there is more than one group | 19-23 |
| Bias | 9 | Describe any efforts to address potential sources of bias | 19 |
| Study size | 10 | Explain how the study size was arrived at | Suppl. p2 |
| Quantitative variables | 11 | Explain how quantitative variables were handled in the analyses. If applicable, describe which groupings were chosen and why | 19-23 |
| Statistical methods | 12 | (*a*) Describe all statistical methods, including those used to control for confounding | 19-23 |
|  |  | (*b*) Describe any methods used to examine subgroups and interactions | 19-23, Supplement |
|  |  | (*c*) Explain how missing data were addressed |  |
|  |  | (*d*) If applicable, explain how loss to follow-up was addressed |  |
|  |  | (*e*) Describe any sensitivity analyses | Supplement p3-4 |
| Results | | |  |
| Participants | 13* | (a) Report numbers of individuals at each stage of study—eg numbers potentially eligible, examined for eligibility, confirmed eligible, included in the study, completing follow-up, and analysed | 5; Figure S1 |
|  |  | (b) Give reasons for non-participation at each stage | Figure S1 |
|  |  | (c) Consider use of a flow diagram | Figure S1 |
| Descriptive data | 14* | (a) Give characteristics of study participants (eg demographic, clinical, social) and information on exposures and potential confounders | 5 |
|  |  | (b) Indicate number of participants with missing data for each variable of interest | Tables 1-3 |
|  |  | (c) Summarise follow-up time (eg, average and total amount) | Figure S1 |
| Outcome data | 15* | Report numbers of outcome events or summary measures over time | Figure 2 |
| Main results | 16 | (*a*) Give unadjusted estimates and, if applicable, confounder-adjusted estimates and their precision (eg, 95% confidence interval). Make clear which confounders were adjusted for and why they were included | Tables 1, 2, 3, Table S1 |
|  |  | (*b*) Report category boundaries when continuous variables were categorized | Tables 1, 2, 3, Table S1 |
|  |  | (*c*) If relevant, consider translating estimates of relative risk into absolute risk for a meaningful time period |  |
| Other analyses | 17 | Report other analyses done—eg analyses of subgroups and interactions, and sensitivity analyses | NA |
| Discussion | | |  |
| Key results | 18 | Summarise key results with reference to study objectives | 14 |
| Limitations | 19 | Discuss limitations of the study, taking into account sources of potential bias or imprecision. Discuss both direction and magnitude of any potential bias | 15 |
| Interpretation | 20 | Give a cautious overall interpretation of results considering objectives, limitations, multiplicity of analyses, results from similar studies, and other relevant evidence | 14-17 |
| Generalisability | 21 | Discuss the generalisability (external validity) of the study results | 16 |
| Other information | | |  |
| Funding | 22 | Give the source of funding and the role of the funders for the present study and, if applicable, for the original study on which the present article is based | Submission system |

| **Table S2: Baseline prevalence and predictors of high density gametocyte infections (≥10 gametocytes/microliter) among infections containing gametocytes at baseline** | | | | | | |
| --- | --- | --- | --- | --- | --- | --- |
| **Predictor** | **N gametocyte positive** | **Weighted proportion containing high density gametocytes** | **Bivariate association** | | **Multivariable association** | |
|  |  |  | **OR [95% CI]** | **p-value** | **OR [95% CI]** | **p-value** |
| Site/school^a^ |  |  |  |  |  |  |
| Bvumbwe | 19 | 0.45 | Ref. | 0.071 | **Ref.^f^** | **0.027** |
| Ngowe | 25 | 0.18 | 0.26 [0.06-1.1] |  | **0.07 [0.01-0.49]** |  |
| Maseya | 71 | 0.13 | 0.19 [0.05-0.65] |  | **0.09 [0.02-0.46]** |  |
| Makwhira | 65 | 0.17 | 0.26 [0.07-0.96] |  | **0.11 [0.02-0.56]** |  |
| Season |  |  |  |  |  |  |
| Dry | 77 | 0.01 | **Ref.** | **0.001** | **Ref** | **0.001** |
| Rainy | 103 | 0.34 | **89.1 [10.6-748]** |  | **118 [10.9-1278]** |  |
| Age (per year increase) | 180 | --- ^b^ | 0.93[0.79-1.1] | 0.374 | **0.80 [0.65-0.99]** | **0.044** |
| Sex |  |  |  |  |  |  |
| Male | 92 | 0.13 | Ref. | 0.118 |  |  |
| Female | 88 | 0.27 | 2.3 [0.81-6.4] |  |  |  |
| Hemoglobin |  |  |  |  |  |  |
| Not anemic | 138 | 0.16 | Ref. | 0.270 |  |  |
| Anemic **^c^** | 41 | 0.23 | 1.8 [0.62-5.4] |  |  |  |
| Fever status **^d^** |  |  |  |  |  |  |
| Afebrile | 78 | 0.13 | **Ref.** | **0.047** | **Ref.** | **0.033** |
| Febrile | 102 | 0.24 | **3.0 [1.0-9.0]** |  | **4.0 [1.1-14.2]** |  |
| Recent treatment **^e^** |  |  |  |  |  |  |
| Not treated | 169 | 0.18 | **Ref.** | **0.038** |  |  |
| Treated | 10 | 0.46 | **5.3 [1.1-25.6]** |  |  |  |
| Slept under a ITN |  |  |  |  |  |  |
| No | 102 | 0.17 | Ref. | 0.398 |  |  |
| Yes | 74 | 0.24 | 1.5 [0.57-4.1] |  |  |  |
| All analyzed variables are included in the table, survey weighted linear regression  ITN= insecticide treated net, OR = Odds Ratio, CI = Confidence interval  ^a^ Univariate association for school, all other associations include school  ^b^ Mean age of students with infections containing high density of gametocytes was 10.5 years compare to 11.0 years among those whose infections contained lower density of gametocytes.  ^c^ Hemoglobin <11.0 g/dL  ^d^ Measured temperature ≥37.5ºC at baseline visit or reported in the last two weeks  ^e^ Reported in the last two weeks  ^f^Adjusted OR for Bvumbwe compared to all other schools in the multivariable model is 10.0 [95%CI: 2.2-46], p=0.004 | | | | | | |

| Table S3: Survey weighted longitudinal mixed-effects logistic regression model of gametocyte prevalence with random effects for strata (school and grade) and student within strata | | | |
| --- | --- | --- | --- |
|  | Coefficient | 95% CI | p-value |
| Treatment (at baseline) Untreated | Ref. |  |  |
| Treated | 3.83 | 2.88, 4.78 | <0.001 |
| Among untreated Day 0 | Ref. |  |  |
| Day 7 | 0.35 | -0.44, 1.14 | 0.37 |
| Day 14 | 0.08 | -1.02, 1.18 | 0.88 |
| Day 42 | 0.39 | -0.25, 1.04 | 0.22 |
| Among treated Day 0 | Ref. |  |  |
| Day 7 | -2.98 | -3.64, -2.33 | <0.001 |
| Day 14 | -3.64 | -4.53, -2.76 | <0.001 |
| Day 42 | -3.78 | -4.53, -3.02 | <0.001 |
| Season Rainy | Ref. |  |  |
| Dry | -0.61 | -1.07, -0.16 | 0.010 |
| School Bvumbwe | Ref. |  |  |
| Ngowe | 0.37 | -0.55, 1.29 | 0.41 |
| Maseya | 1.47 | 0.78, 2.16 | <0.001 |
| Makhuwira | 1.84 | 1.24, 2.43 | <0.001 |
| Coefficient of the random effect of strata (school and grade)= 0.44 (95% CI: 0.196, 1.01); Coefficient of the nested random effect of individual within strata = 3.69 (95% CI: 2.11, 6.45)  Model equation: -4.45 + 3.82*treated + 0.35*day7 + 0.08*day14 + 0.39*day42 + -3.33*treated day 7 + -3.73* treated day 14 + -4.18* treated day 42 + -0.61*dry season + 0.37*Ngowe + 1.47*Maseya + 1.84*Makhuwira | | | |

| Table S4: Survey weighted longitudinal mixed-effects linear regression model of gametocyte density (gametocytes per microliter) with random effects for student | | | |
| --- | --- | --- | --- |
|  | Coefficient | 95% CI | p-value |
| Treatment Untreated | Ref. |  |  |
| Treated | 1.60 | 0.85, 1.79 | <0.001 |
| Among untreated Day 0 | Ref. |  |  |
| Day 7 | 1.51 | 0.32, 2.70 | 0.015 |
| Day 14 | 0.24 | -1.14, 1.63 | 0.721 |
| Day 42 | 0.46 | -0.87, 1.79 | 0.480 |
| Among treated. Day 0 | Ref. |  |  |
| Day 7 | -0.53 | -1.68, 0.62 | 0.352 |
| Day 14 | -2.18 | -4.67, 0.31 | 0.084 |
| Day 42 | -2.32 | -3.91, -0.73 | 0.006 |
| Season Rainy | Ref. |  |  |
| Dry | -2.18 | -2.86, -1.50 | <0.001 |
| School Bvumbwe | Ref. |  |  |
| Ngowe | -0.31 | -1.44, 0.83 | 0.580 |
| Maseya | -0.34 | -1.53, 0.85 | 0.562 |
| Makhuwira | -0.88 | -2.15, 0.39 | 0.167 |
| Coefficient of the random effect of strata = 3.74 (95% CI:2.83, 4.93)  Models including a random effect for strata did not converge.  Model equation: -0.76 + 1.60*treated + 1.51*day7 + 0.24*day14 + 0.46*day42 + -2.04*treated day 7 + -2.42* treated day 14 + -2.78* treated day 42 + -2.18*dry season + -0.31*Ngowe + -0.34*Maseya + -0.88*Makhuwira | | | |

**Figure S2: Age distribution of gametocyte-containing infections in communities surrounding schools**

Color designates age group: school-age children (6-15y) – black; younger children (6-71m) – light grey; adults (>15y) – dark grey.

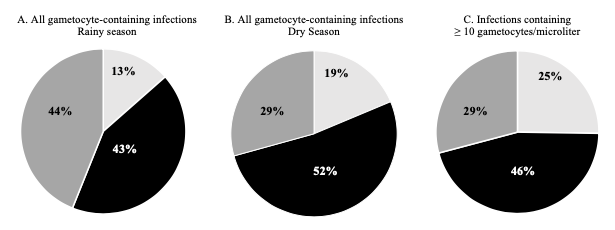

**Text S4: Contribution of school-age children to the population of gametocytes in the communities surrounding schools**

In concurrent household-based surveys in communities surrounding each school, 46% (229/494) of gametocyte-containing infections were found in school-age children, who made up only 35% of the population (Figure S2A and S2B). The predominance of gametocyte-containing infections in school-age children compared to other age groups was consistent between seasons, and across transmission settings, with 55%, 44%, 46%, and 46% occurring in Bvumbwe, Ngowe, Maseya, and Makhuwira, respectively.

The geometric mean gametocyte density was higher in young children (1.45 gametocytes/µl) compared to school-age children (1.05 gametocytes/µl, p_unadjusted_=0.001) and adults (0.56 gametocytes/µl, p_unadjusted_<0.001). However, among the 103 infections with ≥ 10 gametocytes/µl, 47 (46%) were in school age children compared to 26 (25%) in younger children and 30 (29%) in adults (Figure S2C).
